# Increased Association Between Obstructive Sleep Apnea with Hypertension, Diabetes, Dyslipidemia, Chronic Renal Disease, and increasing weight with lower prevalence in cachexia

**DOI:** 10.64898/2025.12.29.25343183

**Authors:** Manouchehr Nasrollahzadeh Saravi, Mohammad Reza Movahed, Mehtrash Hashemzadeh, Ritika Ravindran, Aria Coe, Mehrnoosh Hashemzadeh

**Author notes:** Correspondence: Mehrnoosh Hashemzadeh; 6119 N. Pinchot Rd; Tucson, AZ 85750. Fundings: None. Conflict of interest: None.

## Abstract

Obstructive sleep apnea (OSA), defined as recurrent episodes of upper airway obstruction during sleep, has been associated with multiple cardiovascular and metabolic comorbidities. While prior studies have demonstrated a link between OSA and adverse cardiovascular outcomes, the strength and nature of this relationship remain controversial. We aimed to investigate the association between OSA and major cardiovascular and metabolic conditions, including myocardial infarction (MI), hypertension (HTN), diabetes mellitus (DM), dyslipidemia, obesity, and chronic kidney disease (CKD), as well as the impact of OSA on in-hospital mortality using a large national dataset.

**Materials and Methods:** We conducted a retrospective analysis using the National Inpatient Sample (NIS) database from 2016 to 2020, including over 148 million hospital discharge records. Diagnoses were identified using ICD-10 codes. Multivariate logistic regression was performed to assess adjusted odds ratios (aORs) for each outcome, accounting for potential confounders. A separate mortality analysis was conducted to evaluate in-hospital mortality among patients with and without OSA.

After adjusting for confounding factors, OSA was not independently associated with an increased risk of MI (STEMI or NSTEMI). However, OSA was significantly associated with increased odds of HTN (aOR: 1.79), DM (aOR: 1.40), dyslipidemia, obesity (aOR: 3.36), morbid obesity (aOR: 8.25), and CKD, all with *p* < 0.001. In-hospital mortality analysis revealed worse outcomes in patients with cardiovascular comorbidities such as HTN, DM, dyslipidemia, and obesity.

OSA may not be directly associated with MI, but it is strongly linked to its major risk factors. It may exert a cardioprotective effect in the context of STEMI, potentially due to ischemic preconditioning or heightened clinical surveillance. Conversely, OSA appears to worsen in-hospital mortality in patients with chronic cardiometabolic conditions, underscoring the need for early diagnosis and integrated management strategies tailored to comorbid risk profiles.

## INTRODUCTION

Obstructive sleep apnea (OSA) is a common yet often underdiagnosed condition characterized by recurrent episodes of upper airway obstruction during sleep, resulting in apnea or hypopnea and hypercarbia. (1) In recent years, its prevalence has been on the rise, posing a growing public health concern, as OSA is commonly associated with multiple cardiovascular, metabolic, and neurocognitive conditions. (2) OSA affects approximately 17% of women and 22% of men. (3) Key risk factors include male sex, obesity, increased neck circumference, central fat distribution, genetic predisposition, older age, alcohol or sedative use before sleep, and smoking. (4) The current first-line therapy for OSA is continuous positive airway pressure (CPAP) during sleep, which has been shown to significantly improve symptoms including excessive daytime sleepiness and generally improve quality of life for patients with moderate-to-severe OSA. (5)

While some individuals with OSA present with classic symptoms such as loud snoring, frequent nocturnal awakenings, and excessive daytime sleepiness, many remain asymptomatic. In order to confirm an official OSA diagnosis, patients must undergo a full night, in-laboratory, attend polysomnography sleep study; or must receive type III cardiopulmonary sleep recordings. (5) As a result, many patients who do not display the typical symptoms receive delayed diagnosis and treatment. During sleep monitoring in adults, apnea is scored by a drop in 90% or more of airflow signal excursion for 10 or more seconds using an oro nasal thermal sensor, PAP device flow, or another apnea sensor. (6)

OSA contributes to disrupted sleep patterns and sleep fragmentation, which in turn have been linked to mood disorders, anxiety, cognitive dysfunction, and decreased workplace performance, significantly impacting patients’ quality of life. Sleep apnea-induced fatigue has even been shown to increase risk of motor vehicle accidents. Sleep disturbances because of OSA are also thought to be linked with the development of hypertension, as the use of CPAP as OSA treatment has been shown to improve hypertension. (7, 8)

A substantial body of research has investigated the relationship between OSA and a range of comorbidities, particularly cardiovascular conditions such as myocardial infarction (MI), stroke, heart failure (HF), and hypertension (HTN), as well as metabolic syndrome components including diabetes mellitus (DM), dyslipidemia, and obesity. However, results across studies have been somewhat inconsistent. In particular, the impact of OSA on outcomes following MI remains controversial. (1, 9, 10) While some studies suggest that OSA may exert a protective effect on post-MI outcomes, others report that OSA is associated with worse prognoses, including increased mortality and cardiovascular complications. (11–13) Nonetheless, growing evidence supports the role of OSA in exacerbating cardiovascular risk factors. Recurrent hypoxemia, hypercapnia, intrathoracic pressure changes, and sleep arousals associated with OSA contribute to systemic inflammation, oxidative stress, endothelial dysfunction, sympathetic overactivity, hypercoagulability, and metabolic dysregulation. (8)

In the context of MI, repeated episodes of apnea and hypopnea lead to peripheral vasoconstriction and increased sympathetic drive, elevating blood pressure, heart rate, and cardiac output—thereby increasing myocardial oxygen demand. Simultaneously, hypoxemia and increased afterload from intrathoracic pressure shifts reduce oxygen supply, enhancing myocardial ischemia risk. Additionally, oxidative stress and inflammation contribute to endothelial injury and atherogenesis, further promoting cardiovascular disease. (14)

Given the strong association between OSA and cardiovascular risk, in this study we utilized the National Inpatient Sample (NIS) database to examine these relationships on a large, nationally representative scale. We assessed the association between OSA and major cardiovascular and metabolic outcomes, including ST-elevation and non-ST-elevation myocardial infarction (STEMI, NSTEMI), hypertension, diabetes, dyslipidemia, obesity, morbid obesity, and chronic kidney disease (CKD). Additionally, we analyzed all-cause in-hospital mortality to evaluate the impact of OSA on patient outcomes.

## MATERIALS AND METHODS

### Data Source

We utilized the NIS database from 2016-2020 to generate our study population. The NIS database is the largest publicly available all-payer inpatient database in the United States and includes discharge data from all states participating in the Healthcare Cost and Utilization Project (HCUP) and covers more than 97% of the U.S. population. Our search of the database yielded over 148 million hospitalizations during the study period. We then used the International Classification of Diseases, Tenth Revision (ICD-10) Clinical Modification codes to identify patients with the diagnosis of obstructive sleep apnea (ICD-10 code G47.33)^1^.

### Characteristics

Inclusion criteria for this retrospective study were patients aged 18 years and older. Collected demographic data included age, race, gender, median household income, and expected primary payer. Cardiovascular events and patient co-morbidities were identified through ICD-10 codes. These included STEMI, NSTEMI, diabetes, hypertension, hyperlipidemia, CKD, chronic obstructive pulmonary disease (COPD), cachexia, smoking, obesity and morbid obesity. Hospital characteristics such as hospital bed size, hospital location/teaching status, hospital ownership and hospital region were also incorporated.

### Outcomes

The primary outcome was the prevalence of cardiovascular and cardiometabolic co-morbidities. The secondary outcome was in-hospital mortality in relation to OSA status.

### Statistical Analysis

All analyses were conducted using data from adult patients (≥18 years) hospitalized between 2016 and 2020. Continuous variables were expressed as means with standard deviations (SD) or medians with interquartile ranges (IQR), as appropriate. Differences in continuous variables between patients with and without obstructive sleep apnea (OSA) were assessed using independent samples t-tests for normally distributed variables and Mann–Whitney U tests for non-normally distributed variables. Categorical variables were compared using Chi-square tests. Univariate logistic regression models were initially used to estimate unadjusted odds ratios (ORs) with 95% confidence intervals (CIs) for the association between OSA and clinical outcomes, including mortality and comorbidities. Subsequently, multivariate logistic regression analyses were performed to adjust for potential confounders such as age, sex, race, comorbid conditions, hospital characteristics, and socioeconomic factors. All analyses were conducted following the implementation of population discharge weights. All statistical tests were two-sided, with a significance level set at *p < 0.05*. Analyses were conducted using STATA version 17 (StataCorp, College Station, TX).

## RESULTS

### Baseline Characteristics

Our study incorporated over 148 million documented hospitalized patients between 2016-2020 from the US, of which more than 10 million had OSA diagnosis at the time of admission. Patients with OSA were generally male, white, in the +61-year age group (Mean±SD: 64.08±13.46) and obese (all *p values < 0.001*). smoking and chronic obstructive pulmonary disorders were more ubiquitous in OSA patient, and the patients were more prone to be admitted to Urban teaching hospitals. Baseline characteristics are demonstrated in Table 1.

**Table 1.**
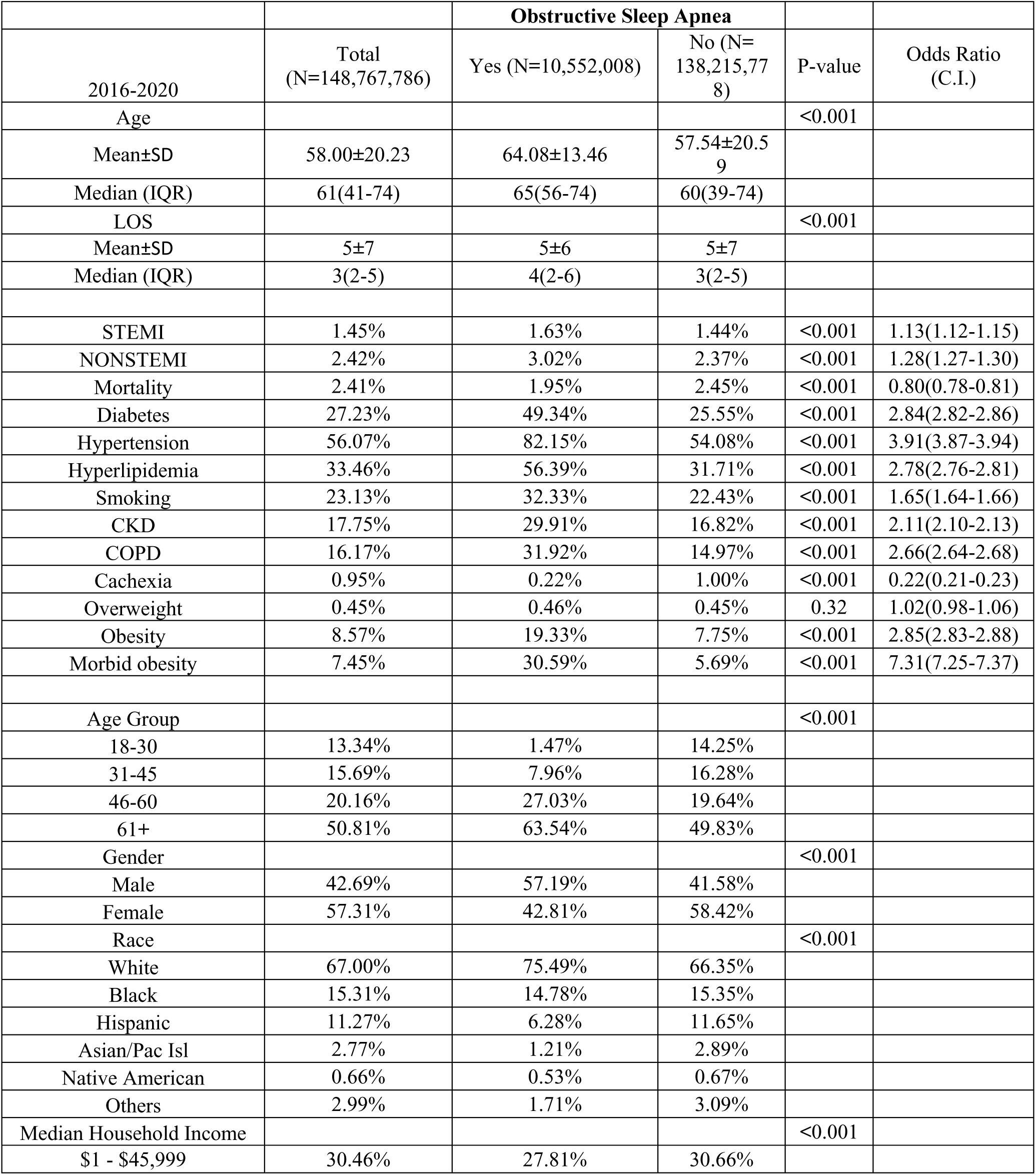
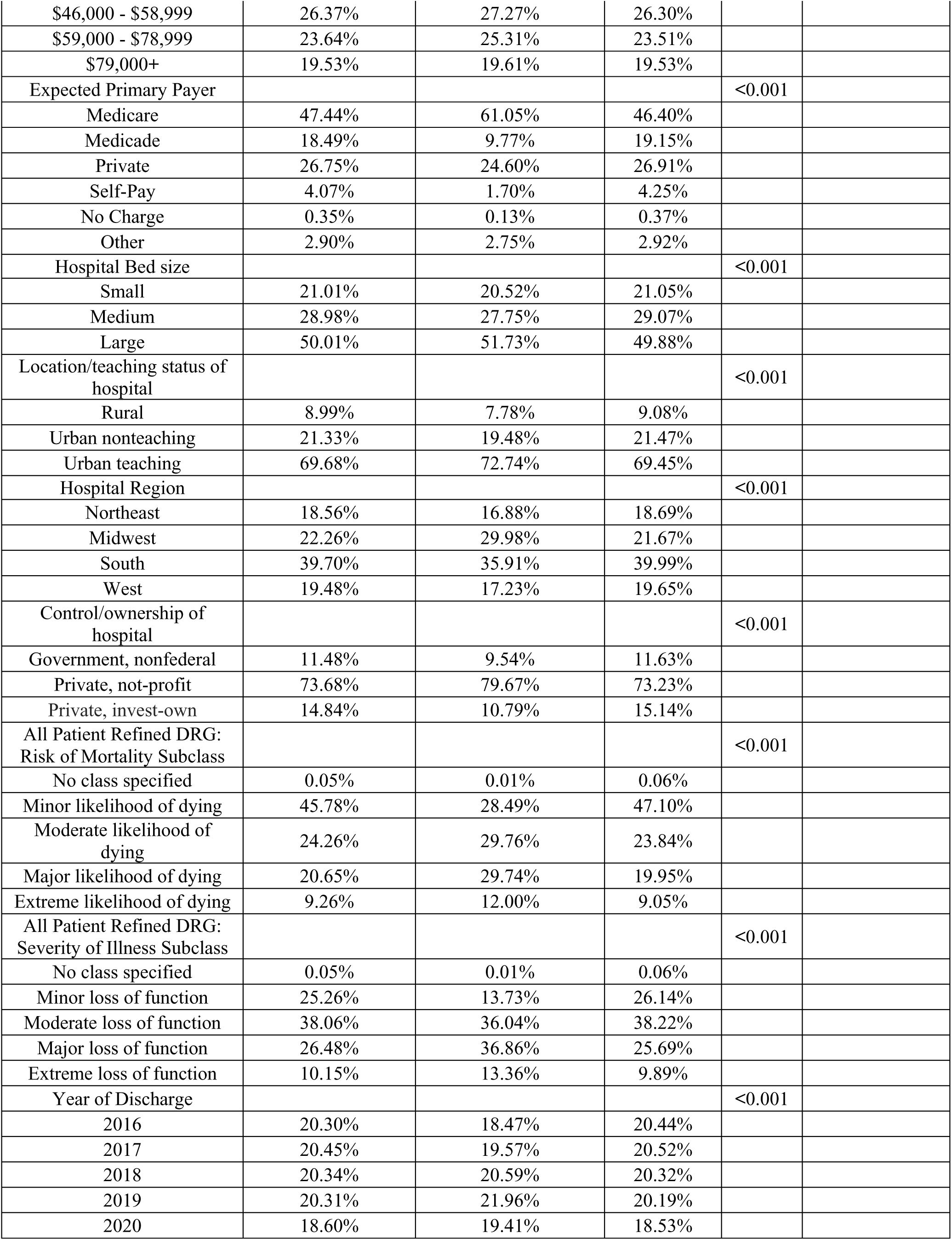
Baseline characteristics and demographic information of subjects.

^1^: Other included codes are uploaded as a supplementary data file

All analyses were conducted following the implementation of population discharge weights.

Inclusion: age>=18

### Multivariate Analysis of OSA and Cardiovascular/Metabolic Conditions

In our findings before adjusting for cofounding variables OSA was associated with MI (both STEMI, NSTEMI), HTN, DM, CKD, obesity and morbid obesity and dyslipidemia (all *p values < 0.001*). however, after conducting multivariate logistic regression analysis to adjust for cofounding variables OSA was negatively associated with MI. (STEMI aOR:0.81, 95 % CI :0.8-0.83, NSTEMI aOR:0.81, 95% CI: 0.8-0.81). on the other hand, OSA was seen to be significantly associated with major metabolic conditions. After multivariate analysis OSA was positively interconnected with HTN (aOR:1.79, 95% CI:1.78-1.81), DM (aOR:1.4, 95% CI:1.39-1.41), Hyperlipidemia (aOR:1.51, 95% CI:1.51-1.52), CKD (aOR:1.19, 95% CI:1.19-1.2). Furthermore, OSA had the strongest independent association with Obesity and morbid obesity (aOR:3.63, 95% CI:3.6-3.67, aOR:8.25, 95% CI:8.18-8.33). On the other hand, as expected OSA and cachexia were negatively associated as OSA patients were 68% less likely to be cachectic (aOR:0.32, 95 % CI:0.31-0.33). more information regarding multivariate analysis is demonstrated in Table 2.

**Figure 1.**
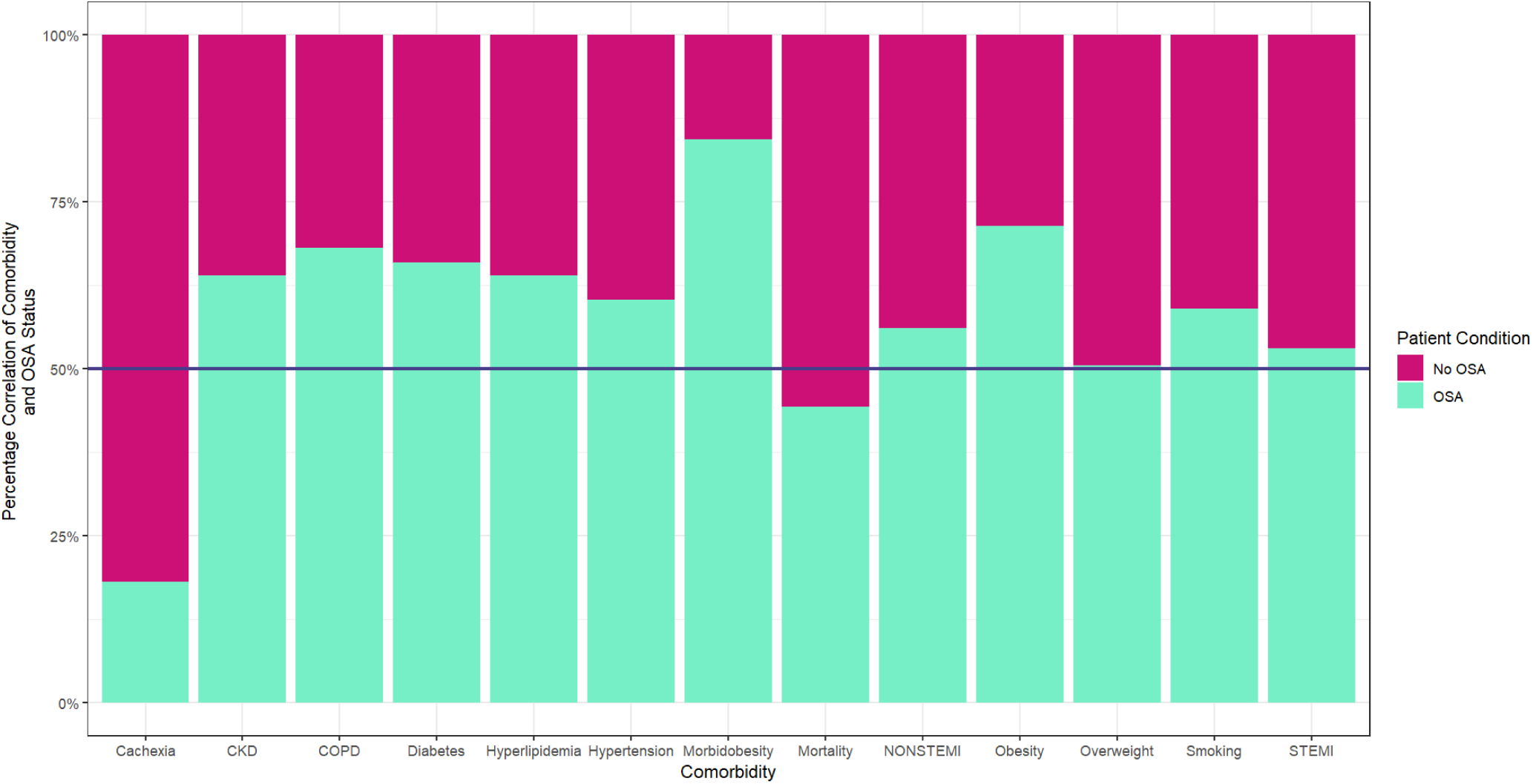
Comparison of different conditions with and without OSA on a percent scale.

**Figure 2.**
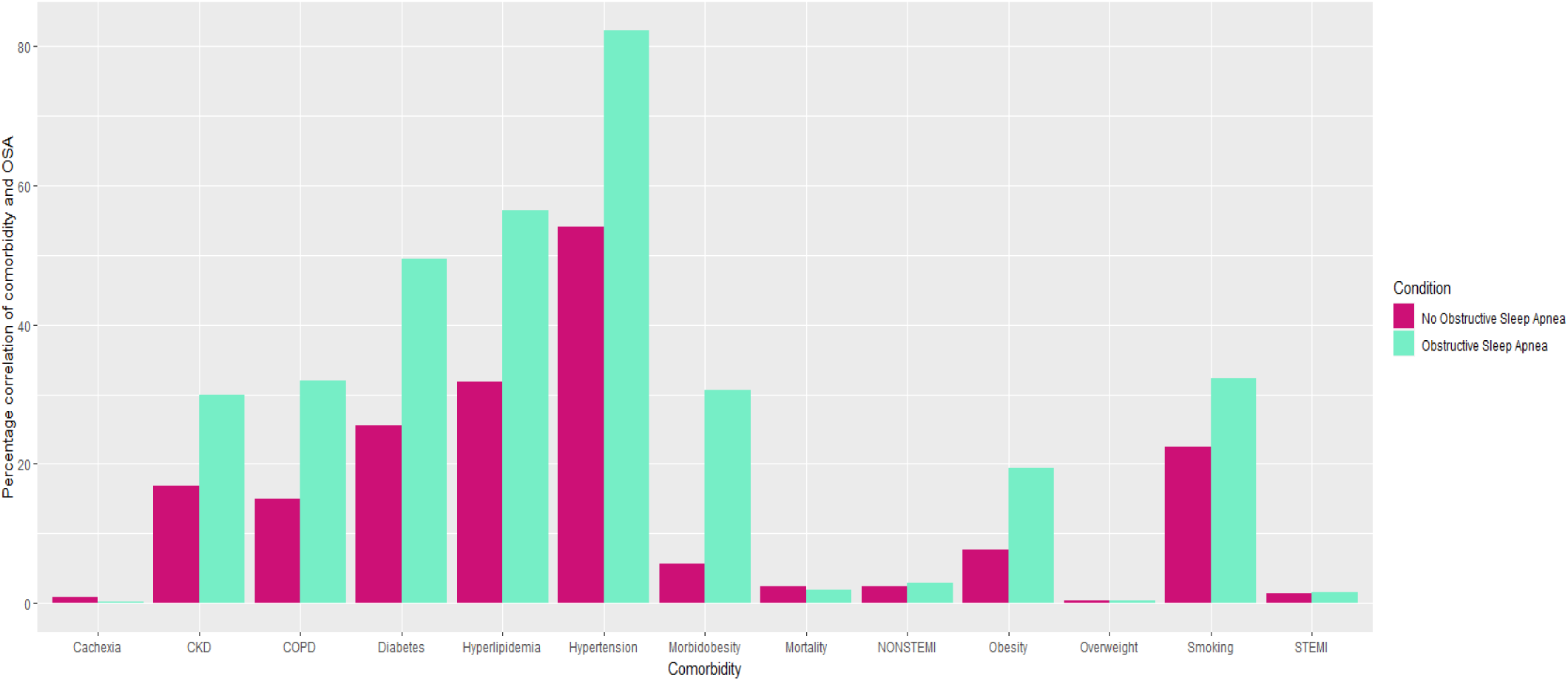
Comparison of different conditions with and without OSA.

**Table 2.** Multivariate analysis of OSA and cardiovascular and cardiometabolic conditions.

| OSA | p-value | OR | 95% C.I.for OR |  |
| --- | --- | --- | --- | --- |
|  |  |  | Lower | Upper |
| STEMI | <0.001 | 0.81 | 0.8 | 0.83 |
| NSTEMI | <0.001 | 0.81 | 0.8 | 0.81 |
| Cachexia | <0.001 | 0.32 | 0.31 | 0.33 |
| Overweight | <0.001 | 1.51 | 1.46 | 1.57 |
| Obesity | <0.001 | 3.63 | 3.6 | 3.67 |
| Morbidobesity | <0.001 | 8.25 | 8.18 | 8.33 |
| Diabetes | <0.001 | 1.4 | 1.39 | 1.41 |
| Hypertension | <0.001 | 1.79 | 1.78 | 1.81 |
| Hyperlipidemia | <0.001 | 1.51 | 1.51 | 1.52 |
| Smoking | <0.001 | 1.15 | 1.14 | 1.15 |
| CKD | <0.001 | 1.19 | 1.19 | 1.2 |
| COPD | <0.001 | 1.86 | 1.85 | 1.87 |

### In-Hospital Mortality Analysis

In-hospital mortality analysis revealed that OSA was not associated with higher mortality in patients with STEMI or NSTEMI compared to those without OSA. Among patients with the following comorbidities—hypertension, diabetes mellitus, dyslipidemia, CKD, COPD, smoking, obesity, and morbid obesity OSA was significantly associated with increased in-hospital mortality (*p* < 0.001 for all). However, there was no significant difference in mortality among overweight patients with and without OSA (*p* = 0.38).

A sex-specific analysis showed that female patients with OSA had lower mortality compared to non-OSA females (37.58% vs. 47.58%, *p* < 0.001), whereas male OSA patients experienced higher mortality than non-OSA males (62.42% vs. 52.44%, *p* < 0.001). Additionally, OSA patients had a shorter length of hospital stay compared to non-OSA patients (LOS: 8 ± 10 vs. 8 ± 12 days). Further details of these mortality findings are presented in Table 3.

**Table 3.** In-hospital mortality analysis of patients with and without OSA.

| Mortality Demographics | Total<br>(N=3,584,119) | Obstructive Sleep Apnea |  | P-value |
| --- | --- | --- | --- | --- |
|  |  | Yes (N=206,090) | No (N=3,378,029) |  |
| Age |  |  |  | <0.001 |
| Mean±SD | 71.21±14.91 | 69.62±12.39 | 71.30±15.05 |  |
| Median (IQR) | 73(62-83) | 71(62-78) | 73(62-83) |  |
| LOS |  |  |  | <0.001 |
| Mean±SD | 8±12 | 8±10 | 8±12 |  |
| Median (IQR) | 4(2-9) | 5(2-11) | 4(1-11) |  |
|  |  |  |  | <0.001 |
| STEMI | 6.37% | 5.80% | 6.40% | <0.001 |
| NONSTEMI | 7.34% | 7.52% | 7.33% | 0.16 |
| Diabetes | 32.72% | 52.14% | 31.53% | <0.001 |
| Hypertension | 65.97% | 82.26% | 64.98% | <0.001 |
| Hyperlipidemia | 33.05% | 50.75% | 31.97% | <0.001 |
| Smoking | 21.42% | 28.22% | 21.01% | <0.001 |
| CKD | 31.26% | 45.36% | 30.40% | <0.001 |
| COPD | 24.94% | 41.08% | 23.96% | <0.001 |
| Cachexia | 4.77% | 1.14% | 4.99% | <0.001 |
| Overweight | 0.20% | 0.21% | 0.19% | 0.38 |
| Obesity | 4.97% | 13.72% | 4.43% | <0.001 |
| Morbidobesity | 6.04% | 26.21% | 4.81% | <0.001 |

## DISCUSSION

In this study, we analyzed over 148 million in-patient records from the United States, of which more than 10 million had a documented diagnosis of OSA at the time of admission. To the best of our knowledge, this represents the largest study to date assessing the relationship between OSA and cardiovascular outcomes using large-scale administrative data.

Our findings indicate that, after adjusting for potential confounding factors, OSA was not associated with an increased risk of either STEMI or NSTEMI. This result contrasts with much of the existing literature, which generally reports an independent association between OSA and MI. (1, 11) Several factors may explain this discrepancy. First, the use of the NIS database limits access to important clinical details, such as OSA severity (typically measured by the apnea–hypopnea index, AHI) and treatment data, including continuous positive airway pressure (CPAP) use. Prior studies have shown that individuals with moderate-to-severe OSA (AHI ≥ 15) are at a significantly higher risk for cardiovascular disease, including MI. (15) Moreover, CPAP therapy has been shown to mitigate cardiovascular risk in OSA patients. (8) It is therefore plausible that a substantial number of patients in our study were either receiving treatment or had less severe disease, potentially attenuating the association between OSA and MI. Second, our analysis was limited to the association between OSA and acute myocardial infarction (STEMI and NSTEMI), whereas OSA has also been strongly linked to other cardiovascular conditions, including heart failure, stroke, and arrhythmias—which were not evaluated in this phase of the study. Third, patients with a known diagnosis of OSA may benefit from enhanced clinical surveillance and early intervention, leading to better management of cardiovascular risk factors. This increased medical attention could contribute to a protective bias, reducing the likelihood of adverse cardiac events being recorded during hospitalization. Additionally, differences in study design, population characteristics, and clinical treatment protocols may also contribute to the variation in findings compared to previous reports. Consistent with our findings, a cohort study involving over 10,000 participants by Kendzerska et al. also reported that, after adjusting for confounding variables, the association between OSA (measured by AHI) and CVD was not statistically significant. (16) The authors stated that this lack of association can be attributed to variability in study methodologies and the differences in diagnostic criteria and definitions of OSA used across studies. Nevertheless, prior research has generally reported a higher prevalence of OSA in patients with acute coronary syndromes, with some studies suggesting rates up to twice as high as in the general population. (14) For example, Kuniyoshi et al., in their study on nocturnal ischemic risk, reported significantly higher rates of MI among patients with OSA compared to those without (32% vs. 7%), further highlighting the interconnected relationship between OSA and cardiovascular events. (17) These conflicting results underscore the complexity of this association and the need for further prospective studies with detailed clinical, diagnostic, and treatment data to fully understand the impact of OSA on acute coronary outcomes.

We found no increased mortality in patients with sleep apnea suffering from myocardial infarction. Some studies suggest that OSA may confer a protective effect through ischemic preconditioning, where recurrent exposure to intermittent hypoxia (IH) induces adaptive responses that make the myocardium more resistant to severe ischemic episodes. (18) Shah et al. found that patients with OSA had lower peak troponin levels, suggesting less severe ischemic injury during acute MI compared to non-OSA patients. Interestingly, patients with more severe OSA (higher AHI) in their cohort had even milder infarctions. (19) Similarly, Borczynski et al. reported lower peak troponin levels in NSTEMI patients with OSA. (13) Another study using ICD-9-coded NIS data reported in-hospital mortality rates of 3.7% in OSA patients compared to 7.4% in non-OSA patients (*p* < 0.001)—a trend that mirrors our findings for STEMI patients. Although that study noted a higher prevalence of STEMI in the OSA group, the post-MI outcomes were more favorable, which the authors attributed to greater diagnostic vigilance, more aggressive management, and possibly ischemic preconditioning. An additional hypothesis is the obesity paradox, whereby obese patients with cardiovascular disease may experience better outcomes due to factors such as earlier symptom onset, protective neurohormonal profiles, and more aggressive treatment regimens. (11) In contrast, Barbé et al. found worse outcomes in OSA patients during acute coronary syndrome (ACS), including higher troponin levels and lower ejection fractions. They attributed this to classical OSA-related mechanisms such as elevated sympathetic tone, systemic inflammation, oxidative stress, endothelial dysfunction, hypercoagulability, and cardiac excitability. (20) The obesity paradox in STEMI patients is well documented. We have shown that OSA patients suffer more from obesity, which is protective in patients presenting with myocardial infarction, as another explanation for the lack of harm in patients with OSA presenting with myocardial infarction. (21) These conflicting findings in the literature underscore the need for future well-controlled, prospective studies to fully elucidate the complex relationship between OSA and myocardial infarction outcomes. Contrary to our findings suggesting a possible cardioprotective role of OSA in STEMI-related in-hospital mortality, our analysis demonstrated worse in-hospital mortality outcomes among OSA patients with other comorbid conditions, including HTN, DM, dyslipidemia, CKD, obesity, and morbid obesity (*p* < 0.05). These results underscore the complex and heterogeneous impact of OSA, which may vary depending on the specific underlying comorbidities. This highlights the critical importance of early diagnosis and proactive management of OSA, particularly in patients with multiple comorbidities, in order to reduce the risk of adverse clinical outcomes in this high-risk population.

In contrast to our findings on MI, the association between OSA and HTN in our study aligns closely with the existing body of literature. The link between OSA and HTN is well established, particularly in cases of resistant or refractory hypertension. (22, 23) While a few studies have reported that the association weakens after adjusting for confounding factors (24)—especially body mass index (BMI)—our multivariate analysis revealed that OSA was independently associated with a significantly higher risk of hypertension (aOR: 1.79; 95% CI: 1.78–1.81). This finding reinforces the biological plausibility and clinical relevance of the relationship and is consistent with a large meta-analysis involving over 51,000 subjects. (25) That analysis reported a progressive increase in hypertension risk with OSA severity: pooled OR = 1.184 (95% CI: 1.093–1.274) for mild OSA, 1.316 (95% CI: 1.197–1.433) for moderate OSA, and 1.561 (95% CI: 1.287–1.835) for severe OSA. Furthermore, a significant association was found between OSA and resistant hypertension, with a pooled OR of 2.842 (95% CI: 1.703–3.980).

Mechanistically, OSA contributes to sustained hypertension through multiple interrelated pathways. Sympathetic nervous system activity, which is normally reduced during sleep, remains persistently elevated in OSA patients. This heightened sympathetic tone carries over into waking hours, leading to chronic vasoconstriction, vascular remodeling, and increased systemic vascular resistance. In parallel, sympathetic activation stimulates the renin–angiotensin–aldosterone system (RAAS), promoting sodium and water retention, which further elevates blood pressure. Additionally, OSA induces systemic inflammation, as evidenced by increased circulating levels of C-reactive protein (CRP), interleukin-6, interleukin-8, and tumor necrosis factor-alpha (TNF-α)—all of which are known to contribute to vascular dysfunction and elevated blood pressure. (8, 26) Treatment with CPAP has been shown to reduce blood pressure, with effects more pronounced in patients with severe OSA. Studies report average reductions of 6–7 mmHg in systolic and 4–5 mmHg in diastolic pressure following CPAP therapy. (27) However, CPAP has not yet demonstrated consistent efficacy in preventing the onset of hypertension in normotensive individuals with OSA, highlighting the need for further research into preventive strategies. (28)

Obesity and morbid obesity showed the strongest associations with OSA after adjusting for confounding variables, with aOR of 3.36 and 8.25, respectively. Conversely, and as anticipated, OSA was negatively associated with cachexia (aOR: 0.32, 95% CI: 0.31–0.33), reflecting the predominance of an excess weight pattern among individuals diagnosed with OSA. These findings highlight the well-established bidirectional relationship between OSA and obesity. Obesity is a recognized risk factor for the development of OSA due to increased upper airway resistance and fat deposition (29), while OSA itself has been linked to weight gain through mechanisms such as sleep fragmentation, hormonal dysregulation, and reduced physical activity (26), further perpetuating the cycle of metabolic dysfunction. A longitudinal study involving normal-weight participants demonstrated that a 10% increase in body weight was associated with a sixfold increase in the risk of developing OSA. Conversely, a 10% reduction in weight led to a 26% decrease in the AHI, indicating a dose-dependent relationship between body weight and OSA severity. (30) Reflecting this relationship, the American Thoracic Society recommends calorie-restrictive dietary interventions for OSA patients with a BMI greater than 25 kg/m^2^. (31)

OSA and obesity together contribute to the synergistic activation of multiple pathophysiological pathways, including sympathetic nervous system overactivity, RAAS activation, endothelial dysfunction, chronic systemic inflammation, and metabolic dysregulation. These interrelated mechanisms significantly elevate the risk of cardiovascular diseases, particularly hypertension. Importantly, studies have shown that combined treatment with CPAP therapy and weight reduction may result in superior cardiovascular outcomes, highlighting the need for a comprehensive and multidisciplinary approach to management in this patient population. (26)

Dyslipidemia is another comorbidity frequently linked to OSA in prior research. (32, 33) Consistent with these findings, our study demonstrated that OSA is independently associated with an increased risk of dyslipidemia, even after adjusting for potential confounding factors. This aligns with a meta-regression analysis by Nadeem et al., which included data from over 18,000 patients and reported a significant association between OSA and elevated levels of total cholesterol, triglycerides (TG), and low-density lipoprotein (LDL)—independent of obesity. (34) Their analysis highlighted LDL, in particular, as a key contributor to the increased cardiovascular risk observed in OSA patients, given its well-established role in the pathogenesis of coronary artery disease and myocardial infarction. (35) Additional studies by Popadić et al. and Xia et al. further support the link between OSA severity and the degree of dyslipidemia, indicating a dose-response relationship. (36, 37) Moreover, given that individuals with OSA are more likely to be obese, this comorbidity may further exacerbate lipid abnormalities, compounding the risk of cardiovascular and cerebrovascular disease. Several pathophysiological mechanisms have been proposed to explain the connection between OSA and dyslipidemia. Intermittent hypoxia (IH)—a hallmark of OSA characterized by repeated cycles of oxygen desaturation and reoxygenation—induces oxidative stress and systemic inflammation. These processes impair lipid clearance, promote lipid synthesis, and lead to the release of pro-inflammatory cytokines, particularly IL-1, which has been shown to alter lipid metabolism in vascular endothelial cells. In addition, sleep fragmentation alone has been associated with elevated LDL-C levels, further contributing to cardiovascular risk. (34, 36, 38, 39) The role of treatment, particularly CPAP, in modifying lipid profiles remains an area of ongoing debate. While some studies have suggested improvements in lipid parameters with CPAP therapy, clinical trials have yet to conclusively establish its impact on dyslipidemia. (40, 41) This underscores the need for future research to determine whether effective OSA treatment can yield meaningful improvements in lipid metabolism. Nevertheless, the high prevalence of dyslipidemia among OSA patients highlights the importance of targeted screening and metabolic monitoring in this population to mitigate the risk of cardiovascular complications.

IH and sleep fragmentation—hallmarks of OSA—are well known to impair glucose metabolism and reduce insulin sensitivity, thereby increasing the risk of developing type 2 DM. These disturbances lead to sympathetic nervous system activation and oxidative stress, both of which further contribute to insulin resistance and metabolic dysregulation. (42) In our study, after adjusting for confounding variables, OSA was independently associated with an increased risk of diabetes mellitus, with an aOR of 1.40 (95% CI: 1.39–1.41). This finding is consistent with prior research, including a meta-analysis involving over 65,000 patients, which reported a 35% increased risk of incident diabetes in individuals with OSA (aOR: 1.35, 95% CI: 1.24–1.47) after adjustment for key covariates. Notably, that analysis also emphasized a bidirectional relationship between OSA and DM. While OSA is a recognized risk factor for the development of diabetes, emerging evidence suggests that diabetes may itself increase the risk of developing OSA, possibly due to autonomic dysfunction, microvascular damage, or obesity-related pathways. (43) This bidirectional link underscores the importance of comprehensive screening and management strategies aimed at early detection and prevention of both conditions. Although the association between OSA and DM is well established, the impact of CPAP therapy on glycemic control and diabetes risk remains uncertain. Some studies suggest modest improvements in insulin sensitivity with CPAP use, while others report no significant effect, especially in patients with poor adherence. (43) These mixed findings highlight the need for larger, well-designed trials to better understand the therapeutic role of OSA treatment in preventing or managing type 2 diabetes.

As previously discussed, IH associated with OSA leads to the generation of ROS, which contribute to endothelial dysfunction and microvascular damage. In addition, elevated systemic inflammation, persistent sympathetic activation, and hypertension—all of which are common in OSA—further exacerbate vascular injury. These vascular changes may impair renal perfusion and contribute to the development of CKD. (43, 44) Although kidney disease may not receive as much attention as other OSA-related comorbidities, it is a clinically significant consequence with the potential to substantially impact quality of life and long-term outcomes. In our study, after adjusting for confounding variables, OSA was independently associated with CKD, a finding that aligns with prior evidence, including a meta-analysis reporting a higher prevalence of CKD among individuals with OSA. (44) Importantly, similar to DM, the relationship between OSA and CKD appears to be bidirectional. CKD may contribute to the development or worsening of OSA through mechanisms such as fluid retention, which can lead to pharyngeal narrowing during sleep, and the accumulation of uremic toxins, which may disrupt central respiratory regulation. (44, 45) This reciprocal relationship highlights the need for increased clinical awareness and screening for renal dysfunction in patients with OSA, as well as the evaluation of sleep disorders in individuals with existing CKD.

Our study has several limitations that should be considered. First, we relied on ICD-10 codes to identify OSA, which may have resulted in misclassification or underdiagnosis, as some patients with undiagnosed or undocumented OSA may have been excluded. Second, due to the retrospective nature of our study and the limitations of the NIS database, we lacked access to data on OSA severity (e.g., AHI) and treatment status, including the use and adherence to CPAP therapy, both of which are known to significantly influence cardiovascular outcomes. Third, our analysis was restricted to in-hospital events and did not include data on long-term outcomes or post-discharge morbidity and mortality. Therefore, the apparent protective association between OSA and in-hospital STEMI-related mortality may not extend beyond the index hospitalization. Fourth, although we used multivariate logistic regression to adjust for a wide range of confounders, the possibility of residual or unmeasured confounding cannot be entirely ruled out. Lastly, while we focused on MI, we did not evaluate other important cardiovascular outcomes such as stroke, heart failure, or arrhythmias, which are also relevant in the OSA population and warrant future investigation. Nonetheless, the large sample size and national representativeness of our dataset provide substantial statistical power and generalizability, which may help mitigate some of these limitations.

## CONCLUSION

OSA, characterized by recurrent episodes of upper airway obstruction during sleep leading to apnea or hypopnea, has been associated with a wide range of cardiometabolic conditions, including MI, HTN, DM, and dyslipidemia. In this large-scale study, we utilized the NIS database and ICD-10 codes to investigate these associations. While our multivariate analysis did not identify an independent association between OSA and acute MI (STEMI or NSTEMI), OSA was significantly associated with key cardiovascular risk factors—including HTN, DM, obesity, and dyslipidemia. This suggests that OSA may contribute indirectly to cardiovascular morbidity by exacerbating underlying risk factors. Moreover, our analysis of in-hospital mortality revealed a potential cardioprotective effect of OSA in patients with STEMI, possibly attributable to mechanisms such as ischemic preconditioning, enhanced clinical surveillance, and more aggressive management in diagnosed individuals. In sum, although OSA may not be directly linked with MI in all populations, it is clearly associated with a higher burden of cardiovascular and metabolic comorbidities. These findings highlight the need for rigorous screening and comprehensive management of OSA patients, as well as the importance of future prospective, controlled studies to better clarify the relationships and optimize prevention strategies in this high-risk group.

## Data availability statement

NIS database is publicly available

